# Pediatric SARS-CoV-2 seroprevalence in Arkansas over the first year of the COVID-19 pandemic

**DOI:** 10.1101/2021.08.04.21261592

**Authors:** Karl W. Boehme, Joshua L. Kennedy, Jessica Snowden, Shana M. Owens, Marianne Kouassi, Ryan L. Mann, Amairani Paredes, Claire Putt, Laura James, Jing Jin, Ruofei Du, Catherine Kirkpatrick, Zeel Modi, Katherine Caid, Namvar Zohoori, Atul Kothari, Bobby L. Boyanton, J. Craig Forrest

**Affiliations:** Department of Microbiology & Immunology, College of Medicine, University of Arkansas for Medical Sciences, Little Rock, AR; Center for Microbial Pathogenesis and Host Inflammatory Responses, University of Arkansas for Medical Sciences, Little Rock, AR; Winthrop P. Rockefeller Cancer Institute, University of Arkansas for Medical Sciences, Little Rock, AR; Department of Pediatrics, College of Medicine, University of Arkansas for Medical Sciences, Little Rock, AR; Department of Internal Medicine, College of Medicine, University of Arkansas for Medical Sciences, Little Rock, AR; Arkansas Children’s Research Institute, Little Rock, AR; Department of Biostatistics, College of Public Health, University of Arkansas for Medical Sciences, Little Rock, AR; Arkansas Department of Health, Little Rock, AR; Department of Bioinformatics, College of Medicine, University of Arkansas for Medical Sciences, Little Rock, AR; Departments of Pathology, Arkansas Children’s Hospital and University of Arkansas for Medical Sciences, Little Rock, AR

**Author notes:** **Corresponding authors:** Karl W. Boehme, Mailing address: 4301 W. Markham Dr. #511, Little Rock, AR 72205, J. Craig Forrest, Mailing address: 4301 W. Markham Dr. #511, Little Rock, AR 72205, Phone: (501), Joshua L. Kennedy, Mailing address: 13 Children’s Way, Slot 512-13, Little Rock, AR 72205. These authors contributed equally to the work (Karl W. Boehme, Joshua L. Kennedy). **Funding:** We acknowledge funding from the state of Arkansas through the Coronavirus Aid, Relief, and Economic Security (CARES) Act, the UAMS Time-Sensitive COVID-10 Research Award Program, the UAMS Translational Research Institute (UL1TR000039, TL1TR003109, and UL1TR003107), and the Center for Microbial Pathogenesis and Host Inflammatory Response (P20 GM103625). The funders had no role in the study design, data collection, and interpretation.

**Keywords:** COVID-19, SARS-CoV2, serology, serum, antibody, child, epidemiology

## Abstract

**Background:** SARS-CoV-2 seroprevalence studies have largely focused on adults but little is known about spread in children. We determined SARS-CoV-2 seroprevalence in children and adolescents from Arkansas over the first year of the COVID-19 pandemic.

**Methods:** We tested remnant serum samples from children from 1–18 years who visited Arkansas hospitals or clinics for non-COVID19-related reasons from April, 2020 through April, 2021 for SARS-CoV-2 antibodies. We used univariable and multivariable regression models to determine association between seropositivity and participant characteristics.

**Results:** Among 2400 participants, seroprevalence rose from 7.9% in April/May 2020 (95% CI, 4.9-10.9%) to 25.8% in April 2021 (95% CI, 22.2-29.3%). Hispanic and black children had a significantly higher association with antibody positivity than white children in multiple sampling periods.

**Conclusions:** By spring 2021, most children in Arkansas had not been infected with SARS-CoV-2. With the emergence of SARS-CoV-2 variants, recognition of long-term effects of COVID-19, and the lack of an authorized pediatric SARS-CoV-2 vaccine, these results highlight the importance of including children in SARS-CoV-2 public health, clinical care, and research strategies. These findings are important for state and local officials as they consider measures to limit SARS-CoV-2 spread in schools and daycares for the 2021–2022 school year.

## INTRODUCTION

Severe acute respiratory syndrome coronavirus-2 (SARS-CoV-2) emerged in late 2019 and spread globally to cause the coronavirus disease of 2019 (COVID-19) pandemic [1, 2]. SARS-CoV-2 has infected nearly 200 million and killed more than 4 million people worldwide, causing massive disruptions to daily life and untold economic losses [3]. SARS-CoV-2 was first detected in the United States in early 2020 and has caused over 600,000 deaths [4-6] Children were thought to be important for driving SARS-CoV-2 transmission [7, 8]. In March 2020, schools nationwide closed for in-person instruction and classes moved online to curtail SARS-CoV-2 spread [7, 8]. However, pediatric infection rates, and thereby the potential level of immunity in children, is not known [9, 10]. Because approval of SARS-CoV-2 vaccines for those 12 years and under is not expected until late 2021, knowing infection rates in children is important information for school officials as they consider risks and protection measures for the 2021–2022 school year.

Children typically experience less severe COVID-19 and may be more likely to have asymptomatic infections, allowing them to unknowingly spread SARS-CoV-2 [11]. With asymptomatic infections estimated as high as 50%, estimating true infection rates remains challenging [12, 13]. Although nucleic acid testing can identify active SARS-CoV-2 cases, most infections clear within two weeks and leave asymptomatic cases undocumented [12-14]. Antibodies generated against prior infections can last for months to years [10, 15]. The presence of SARS-CoV-2 antibodies in the blood can indicate that a person was infected at some point in the pandemic. Serological studies can capture asymptomatic and symptomatic cases, and more accurately estimate of how many people have been infected with SARS-CoV-2 [10, 15]. Here, we present the results of a seroprevalence study of children ages 1–18 years of age who visited hospitals or regional clinics in Arkansas for non-COVID19-related reasons over the first year of the COVID-19 pandemic.

## METHODS

### Human specimens

All human specimens were obtained with oversight from the UAMS Institutional Review Board (IRB), and waiver of consent and HIPAA applied. Remnants of serum samples collected for routine, non-COVID-19-related clinical laboratory tests were obtained from Arkansas Children’s Hospital (ACH, Little Rock, AR), Arkansas Children’s Northwest (Springdale, AR), and UAMS Family Medical Centers (Ft. Smith, AR and Pine Bluff, AR). Samples were de-identified prior to testing. Inclusion criteria were ages 1–18 years and Arkansas resident. Samples were excluded with the following diagnosis codes: immunodeficiency (primary immune deficiency (D80–D89)), transplant recipient (codes beginning with Z94), and cancer (C00–D49). Samples from patients receiving chemotherapy (prior two months), steroids (prior 30 days), and/or intravenous immunoglobulin (prior 6 months) were excluded. Samples were selected by pathologists at each clinic after they were considered remnant. Clinical and demographic variables were stored in a secure REDCap database [16, 17] and included age, sex, race/ethnicity, zip code, and county of residence. Metropolitan status was determined by cross-1.0 (Ref)erencing zip codes with the Federal Office of Rural Health Policy (FORHP) data files identifying non-metropolitan counties and rural census tracts.

### Protein production and purification

HEK293T cells were cultured in Dulbecco’s minimum essential media (DMEM; Gibco) supplemented with 10% heat-inactivated calf serum (CS, VWR), 2 mM L-glutamine (Invitrogen), and 100 U/mL penicillin/100 µg/mL streptomycin (Invitrogen). Briefly, 15 cm dishes seeded with 9×10^6^ cells on the preceding day were transfected with 20 µg of pCAGGS-SARS-CoV-2 Wuhan-Hu-1 RBD-C-terminal 6-His tag (BEI Resources), pCAGGS SARS-CoV-2 Wuhan-Hu-1 ectodomain Spike glycoprotein gene-C-terminal 6-His tag, or pCMV3 2019-nCoV nucleoprotein-C-terminal 6-His tag (Sino Biological) using polyethylamine (PEI) at a 1:3 ratio. DNA:PEI mixtures were incubated at RT for 10 mins and added to cells with the media volume reduced to 5 mL/dish. After 4–6 hours, the media volume was raised to 25 mL. SARS-CoV-2 RBD and Spike ectodomain proteins were purified as described [18]. Nucleoprotein (N) was isolated as described above under denaturing conditions [19]. Protein concentration was measured by DC Protein Assay (BioRad). Purified proteins were confirmed by Coomassie and western blot using antigen-specific antibodies and stored at −80°C.

### ELISAs

Serum was inactivated at 56°C for 1 h and initially screened by ELISA specific for the receptor-binding domain (RBD) of the SARS-CoV-2 S protein [20-26]. Commercial anti-RBD (Sino Biologicals; 1:2500 dilution) and SARS-CoV-2-RT-PCR-positive patient sera (1:50 dilution) served as positive controls. Purified human IgG (Sigma; 1:2500 dilution) and pre-COVID-19 patient sera (1:50 dilution) served as negative controls. Serum IgM and IgG were detected using horse-radish peroxidase-conjugated anti-human IgM+IgG (Jackson ImmunoResearch; diluted 1:5000 in PBS-T + 1% milk) and SureBlue TMB 1-Component Peroxidase Substrate (SeraCare; 75 µL). After 5 min, the reaction was terminated using TMB Stop Solution (SeraCare; 75 µL). The OD_450_ was measured using a FluoStar Omega plate reader (BMG Labtech). The final OD_450_ was calculated by subtracting the mean OD_450_ of blank wells from the mean OD_450_ of duplicate samples. The statistical cutoff for RBD binding was defined as the mean OD_450_ + 3 standard deviations of pre-COVID-19 sera [26].

Confirmation RBD-positive specimens was performed using a Four-Antigen Confirmation Test (FACT) ELISA. An additional 5% of negative sera were randomly selected and tested in parallel. Plates were coated with 2 µg/mL RBD, spike, nucleoprotein, or bovine serum albumin (BSA; Sigma Aldrich) and FACT ELISA was performed as above. Confirmed positive samples were defined by (i) a mean signal for any viral antigen over 0.6 OD_450_, (ii) BSA-subtracted viral antigen value >0.3 OD_450_, (iii) and at least two positive antigens (RBD+/Spike+, RBD+/N+, Spike+/N+, or RBD+/Spike+/N+). The sensitivity and specificity of the assay were 94.6% and 100%, respectively, based on 37 pre-COVID-19 and 19 RT-PCR-confirmed SARS-CoV-2-positive sera.

### Statistical analyses

The descriptive analysis was conducted for the study population demographics by wave. SARS-CoV-2 antibody positivity rates were reported with 95% confidence intervals (CI) using exact binomial distributions. The 2019 U.S. Census Bureau Arkansas state population estimates (age ≤ 18) were used to calculate the standardized positivity rates [27]. The age- and sex-standardized positivity rates were reported for each wave. Univariable and multivariable analyses were performed to examine the relationship between variables and the SARS-CoV-2 antibody positivity rate for each wave Relative risk was estimated by modified Poisson regressions with robust error variance as the measure to characterize association effects [28]. For multivariable analyses, factors being considered were age group, sex, and race/ethnicity. The statistical significance level was set at 0.05. We conducted all analyses using SAS version 9.4 (SAS Institute).

## RESULTS

### Enrollment and demographic representation

We collected 2400 total remnant pediatric serum samples across five collection periods (waves): Wave 1, April 2 – May 6, 2020 (n = 316); Wave 2, June 6 – August 10, 2020, (n = 300); Wave 3, September 8 – October 17, 2020, (n = 594); Wave 4, November 7 – December 17, 2020, (n = 600); and Wave 5, April 5 – April 28, 2021, (n = 590) (Figure 1). Table 1 shows the demographic characteristics for the study. The 1 – 4 years age group had the fewest samples (n = 328, 13.7%) while the 10 – 14 years group had the most samples (n = 795, 33.2%). Females represented 54.8% (n = 1311), whereas males represented 45.2% (n = 1082) of total samples analyzed. The racial/ethnic distribution of the study population was 55.8% non-Hispanic white (n = 1289), 22.5% non-Hispanic black (n = 520), and 15.2% Hispanic (n = 351). A total of 151 specimens (6.5%) were collected from patients that did not identify as white, black, or Hispanic. Most samples were collected from children living in urban (n = 1564, 75.3%) compared to rural areas (n = 513, 24.7%). Obesity was the most common co-morbidity reported (n = 337, 14.1%), followed by asthma (n = 239, 10.0%), diabetes mellitus (n = 113, 4.7%), and hypertension (n = 111, 4.6%) (Supplemental table 1).

**Table 1.** Sample population demographics for SARS-CoV-2 antibody testing in Arkansas from April 2, 2020 to April 28, 2021.

| Number (%)<sup>a</sup> |  |  |  |  |  |  |
| --- | --- | --- | --- | --- | --- | --- |
| Characteristic | Wave 1<br>(n = 316) | Wave 2<br>(n = 300) | Wave 3<br>(n = 594) | Wave 4<br>(n = 600) | Wave 5<br>(n = 590) | Overall<br>(N = 2400) |
| Collection dates | April 2 –<br>May 6, 2020 | June 6 –<br>August 10, 2020 | September 8 –<br>October 17, 2020 | November 7 –<br>December 17, 2020 | April 5 –<br>April 28, 2021 | April 2, 2020 –<br>April 28, 2021 |
| Age category |  |  |  |  |  |  |
| 1-4 years | 56 (17.7) | 33 (11) | 81 (13.6) | 83 (13.9) | 75 (12.8) | 328 (13.7) |
| 5-9 years | 61 (19.3) | 75 (25) | 112 (18.9) | 116 (19.5) | 142 (24.2) | 506 (21.1) |
| 10-14 years | 90 (28.5) | 98 (32.7) | 205 (34.5) | 214 (36.0) | 188 (32) | 795 (33.2) |
| 15-18 years | 109 (34.5) | 94 (31.3) | 196 (33) | 182 (30.6) | 183 (31.1) | 764 (31.9) |
| Sex |  |  |  |  |  |  |
| Female | 161 (50.9) | 155 (51.7) | 324 (54.5) | 338 (56.8) | 333 (56.5) | 1311 (54.8) |
| Male | 155 (49.1) | 145 (48.3) | 270 (45.5) | 257 (43.2) | 255 (43.2) | 1082 (45.2) |
| Race/Ethnicity |  |  |  |  |  |  |
| White | 178 (56.5) | 154 (52.2) | 327 (57.2) | 312 (54.5) | 318 (57.2) | 1289 (55.8) |
| Black | 91 (28.9) | 53 (18.0) | 117 (20.5) | 150 (26.2) | 109 (19.6) | 520 (22.5) |
| Hispanic | 28 (8.9) | 64 (21.7) | 95 (16.6) | 77 (13.4) | 87 (15.7) | 351 (15.2) |
| Other | 18 (5.7) | 24 (8.1) | 33 (5.8) | 34 (5.9) | 42 (7.6) | 151 (6.5) |
| Metropolitan status<sup>b</sup> |  |  |  |  |  |  |
| Urban | Not Reported | 251 (83.7) | 458 (77.1) | 424 (71.3) | 431 (73.3) | 1564 (75.3) |
| Rural | Not Reported | 49 (16.3) | 136 (22.9) | 171 (28.7) | 157 (26.7) | 513 (24.7) |

<sup>a</sup>Percentages calculated from non-missing values for all periods. Missing values in Wave 1 included race/ethnicity (n = 1) and metropolitan status (n = 316). Missing values in Wave 2 included race/ethnicity (n = 5). Missing values in Wave 3 included race/ethnicity (n = 22). Missing values in Wave 4 included age category (n = 5), sex (n = 5), race/ethnicity (n = 27), and metropolitan status (n = 5). Missing values in Wave 5 included age category (n = 4), sex (n = 2), race/ethnicity (n = 34), and metropolitan status (n = 2).

**Figure 1.**
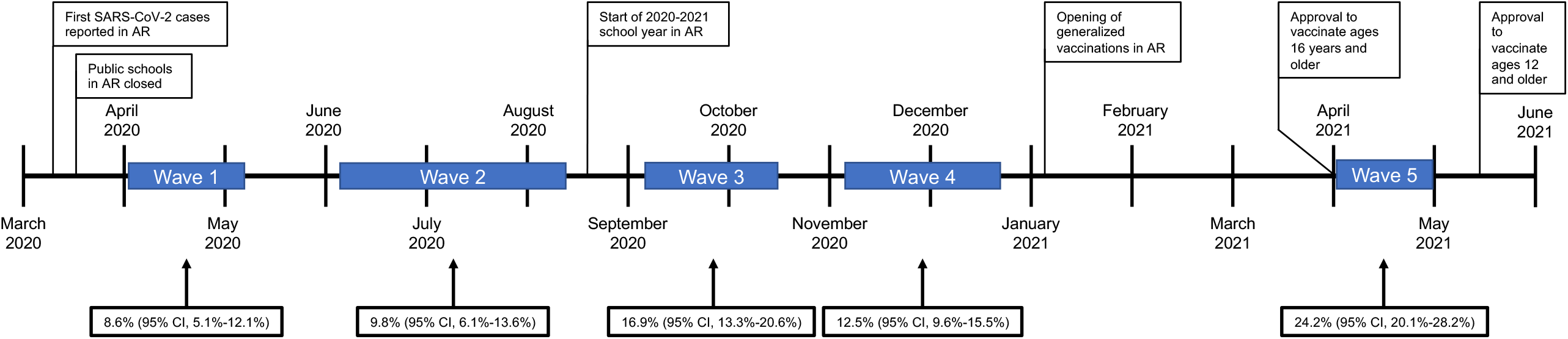
Timeline of Arkansas COVID-19 milestones and sample collection waves. The schematic shows the timeline for relevant study-related events. The sample collection periods (waves) are indicated by blue boxes. The age- and sex-standardized seroprevalence rate for each wave is indicated as the percent (95% CI).

### Estimates of seroprevalence

Table 2 shows the SARS-CoV-2 seroprevalence rates by age, sex, and race/ethnicity, and the age- and sex-standardized estimates. The antibody positivity rate for Wave 1 was 7.9% (95% CI, 4.9%-10.9%), which increased in Wave 2 to 9.7% (95% CI, 6.3%-13.0%) and Wave 3 to 16.2% (95% CI, 13.2%-19.1%), followed by a slight decrease to 13.2% (95% CI, 10.5%-15.9%) in Wave 4. The seroprevalence rate was the highest in Wave 5 at 25.8% (95% CI, 22.2%-29.3%). The overall trend was statistically significant (p<0.0001 using the Cochran-Armitage trend test). When standardized to match Arkansas population demographics [27], seroprevalence rates followed a similar trend to the non-adjusted rates, increasing over Waves 1, 2, and 3, with a decrease in Wave 4 and a peak of 24.2% (95% CI, 22.2%-29.3%) in Wave 5 (Figure 2). The 1 – 4 year-old age group had the highest seroprevalence rates in Wave 1 (10.7%), Wave 2 (15.2%), and Wave 3 (19.8%), but the lowest in Wave 4 (7.2%) and Wave 5 (16.0%). The 15 – 18 year-old group had the highest percentage of reactive specimens in Wave 4 (14.8%) and 10 – 14 year-olds were the highest in Wave 5 (30.9%). No statistically-significant difference was observed between males and females.

**Table 2.** Age-specific, sex-specific, race/ethnicity-specific SARS-CoV-2 seroprevalence estimates in Arkansas from April 2, 2020 to April 28, 2021.

| Persons reactive, % (95% CI) (No.) |  |  |  |  |  |
| --- | --- | --- | --- | --- | --- |
| Characteristic | Wave 1<br>(n = 316) | Wave 2<br>(n = 300) | Wave 3<br>(n = 594) | Wave 4<br>(n = 600) | Wave 5<br>(n = 590) |
| Collection dates | April 2 –<br>May 6, 2020 | June 6 –<br>August 10, 2020 | September 8 – October<br>17, 2020 | November 7 – December<br>17, 2020 | April 5 –<br>April 28, 2021 |
| Age category |  |  |  |  |  |
| 1-4 years | 10.7 (2.4-19.1) (6) | 15.2 (2.2-28.1) (5) | 19.8 (10.9-28.6) (16) | 7.2 (1.54-12.9) (6) | 16.0 (7.5-24.5) (12) |
| 5-9 years | 4.9 (0.0-10.5) (3) | 8.0 (1.7, 14.3) (6) | 18.8 (11.4-26.1) (21) | 12.9 (6.7-19.1) (15) | 19.0 (12.5,-25.5) (27) |
| 10-14 years | 8.9 (2.9-14.9) (8) | 11.2 (4.9-17.6) (11) | 14.2 (9.3-19.0) (29) | 14.0 (9.3-18.7) (30) | 30.9 (24.2-37.5) (58) |
| 15-18 years | 7.3 (1.4-12.3) (8) | 7.4 (2.0-12.9) (7) | 15.3 (10.2-20.4) (30) | 14.8 (9.6-20.0) (27) | 29.5 (22.8-36.2) (54) |
| Sex |  |  |  |  |  |
| Female | 8.7 (4.3-13.1) (14) | 9.0 (4.5-13.6) (14) | 17.3 (13.1-21.4) (56) | 13.6 (9.9-17.3) (46) | 27.0 (22.2-31.8) (90) |
| Male | 7.1 (3.0-11.2) (11) | 10.3 (5.3-15.4) (15) | 14.8 (10.6-19.1) (40) | 12.5 (8.4-16.5) (32) | 23.9 (18.7-29.2) (61) |
| Race/Ethnicity |  |  |  |  |  |
| White | 9.0 (4.7-13.2) (16) | 9.7 (5.0-14.5) (15) | 10.7 (7.3-14.1) (35) | 11.5 (8.0-15.1) (36) | 17.9 (13.7-22.2) (57) |
| Black | 5.5 (0.7-10.3) (5) | 3.8 (0.0-9.1) (2) | 17.1 (10.2-24.0) (20) | 15.3 (9.5-21.2) (23) | 28.4 (19.8-37.0) (31) |
| Hispanic | 10.7 (0.0-22.9) (3) | 14.1 (5.3-22.8) (9) | 29.5 (20.1-38.8) (28) | 20.8 (11.5-30.0) (16) | 43.7 (33.0-54.3) (38) |
| Other | 5.6 (0.0 -7.3) (1) | 12.5 (0.0-26.8) (3) | 27.3 (11.2-43.4) (9) | 2.9 (0.0-8.9) (1) | 35.7 (20.6-50.8) (15) |
| Total | 7.9 (4.9-10.9) (25) | 9.7 (6.3-13.0) (29) | 16.2 (13.2-19.1) (96) | 13.2 (10.5-15.9) (79) | 25.8 (22.2-29.3) (152) |

<sup>a</sup>Standardized positivity rates were calculated based on the 2019 U.S. Census Bureau Arkansas state population estimates (age ≤ 18).<sup>27</sup>

**Table 3.**
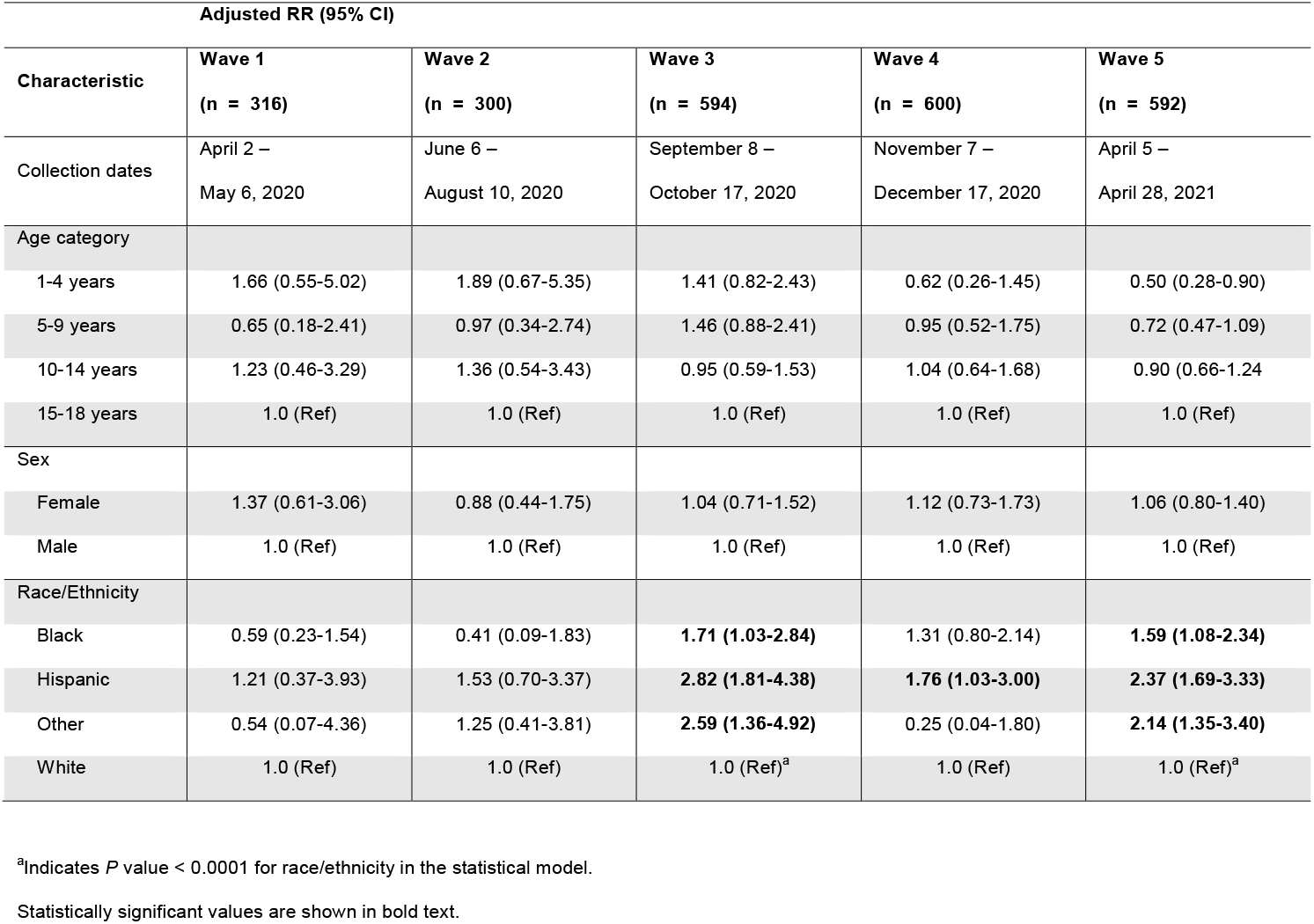
Age- and sex-adjusted association with SARS-CoV-2 antibody positivity by time period.

| Adjusted RR (95% CI) |  |  |  |  |  |
| --- | --- | --- | --- | --- | --- |
| Characteristic | Wave 1<br>(n = 316) | Wave 2<br>(n = 300) | Wave 3<br>(n = 594) | Wave 4<br>(n = 600) | Wave 5<br>(n = 592) |
| Collection dates | April 2 –<br>May 6, 2020 | June 6 –<br>August 10, 2020 | September 8 –<br>October 17, 2020 | November 7 –<br>December 17, 2020 | April 5 –<br>April 28, 2021 |
| Age category |  |  |  |  |  |
| 1-4 years | 1.66 (0.55-5.02) | 1.89 (0.67-5.35) | 1.41 (0.82-2.43) | 0.62 (0.26-1.45) | 0.50 (0.28-0.90) |
| 5-9 years | 0.65 (0.18-2.41) | 0.97 (0.34-2.74) | 1.46 (0.88-2.41) | 0.95 (0.52-1.75) | 0.72 (0.47-1.09) |
| 10-14 years | 1.23 (0.46-3.29) | 1.36 (0.54-3.43) | 0.95 (0.59-1.53) | 1.04 (0.64-1.68) | 0.90 (0.66-1.24) |
| 15-18 years | 1.0 (Ref) | 1.0 (Ref) | 1.0 (Ref) | 1.0 (Ref) | 1.0 (Ref) |
| Sex |  |  |  |  |  |
| Female | 1.37 (0.61-3.06) | 0.88 (0.44-1.75) | 1.04 (0.71-1.52) | 1.12 (0.73-1.73) | 1.06 (0.80-1.40) |
| Male | 1.0 (Ref) | 1.0 (Ref) | 1.0 (Ref) | 1.0 (Ref) | 1.0 (Ref) |
| Race/Ethnicity |  |  |  |  |  |
| Black | 0.59 (0.23-1.54) | 0.41 (0.09-1.83) | **1.71 (1.03-2.84)** | 1.31 (0.80-2.14) | **1.59 (1.08-2.34)** |
| Hispanic | 1.21 (0.37-3.93) | 1.53 (0.70-3.37) | **2.82 (1.81-4.38)** | **1.76 (1.03-3.00)** | **2.37 (1.69-3.33)** |
| Other | 0.54 (0.07-4.36) | 1.25 (0.41-3.81) | **2.59 (1.36-4.92)** | 0.25 (0.04-1.80) | **2.14 (1.35-3.40)** |
| White | 1.0 (Ref) | 1.0 (Ref) | 1.0 (Ref)<sup>a</sup> | 1.0 (Ref) | 1.0 (Ref)<sup>a</sup> |

<sup>a</sup>Indicates *P* value < 0.0001 for race/ethnicity in the statistical model.
Statistically significant values are shown in bold text.

**Figure 2.**
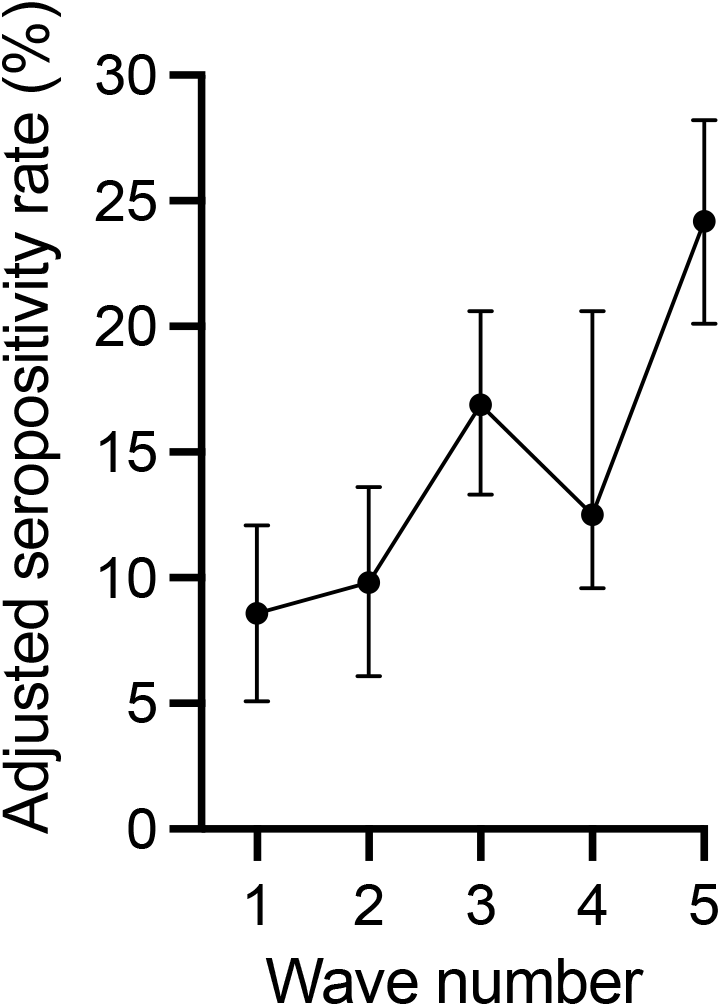
Age- and sex-standardized seroprevalence rates by wave. Error bars indicate 95% CI.

Hispanics had the highest seroprevalence rate in each wave, reaching a maximum of 43.7% in Wave 5. White persons had a higher seroprevalence rate in Waves 1 and 2, but the lowest in Waves 3, 4, and 5. Concomitantly, the antibody reactivity rate in black persons was lower compared to white persons in Waves 1 and 2, but higher in Waves 3, 4, and 5. The highest seroprevalence rates for white (17.9%) and black (28.2%) children was observed in Wave 5. Notably, the peak seroprevalence for Hispanic children was 5.5-fold higher compared to white children and 1.6-fold higher than black children.

There was no significant marginal association between age, sex, or metropolitan status and the likelihood of testing positive for SARS-CoV-2 antibodies (Supplemental table 2). However, Hispanic and black children were more likely to test positive for antibodies than white children during multiple waves after adjusting for age and sex. Hispanic children showed a higher relative risk of testing positive compared to white children in Wave 3 (RR 2.82 95% CI, 1.81-4.38), Wave 4 (RR 1.76 95% CI, 1.03-3.00), and Wave 5 (RR 2.37 95% CI 1.69-3.33). Blacks also showed a higher risk of having antibodies compared to white children in Wave 3 (RR 1.71 95% CI, 1.03-2.84) and Wave 5 (RR 1.59 95% CI, 1.08-2.34). The risk of testing positive for antibodies was higher for children of other races/ethnicities in Wave 3 (RR 2.59 95% CI, 1.36-4.92) and Wave 5 (RR 2.14 95% CI, 1.35-3.40).

Children with asthma (unadjusted RR 4.83 95% CI, 1.96-11.87; *P* = 0.003) or diabetes (unadjusted RR 4.17 95% CI, 1.49-11.67; *P* = 0.007) had higher risk of having antibodies of SARS-CoV-2 than children who did not have asthma or diabetes in Wave 1 (Supplemental table 3). However, this difference was not observed in the remaining waves. PCR testing was performed for 714 of 2400 total specimens, with 38 positive PCR tests reported (Supplemental table 4). A positive RT-PCR test was significantly associated with antibody positivity in Waves 2 through 5 (Supplemental table 5).

## DISCUSSION

Our results demonstrate that by the end of April 2021, approximately 25% of children in Arkansas had SARS-CoV-2-specific antibodies. This percentage is much higher than the total number of confirmed cases, which on April 28, 2021 was 11% for the total population of Arkansas (335,288 positive cases according the Arkansas Department of Health, population of 3,011,524 according to 2019 census data). This finding indicates that those children had been infected with SARS-CoV-2 and are likely to have at least some natural immunity. Conversely, our findings suggest that most children in Arkansas have not been infected with SARS-CoV-2 and remain susceptible to infection. Although COVID-19 has been less severe in children than adults early in the pandemic, emergence of the SARS-CoV-2 delta variant in May 2021 has driven up infection and hospitalization rates, including among those under 18 years of age [11, 29, 30]. Developing multisystem inflammatory syndrome in children (MIS-C), a severe inflammatory disorder that results from a current or recent SARS-CoV-2 infection, is also a risk for those under 18 years [31-33]. Increased SARS-CoV-2 transmission rates combined with a highly susceptible pediatric population raises the possibility that SARS-CoV-2 could spread rapidly in schools and daycares during the upcoming school year. More children infected with SARS-CoV-2 could lead to an increase in the number of severe COVID-19 and MIS-C cases, and a rise in pediatric deaths.

The first SARS-CoV-2 infections in Arkansas were reported in March 2020 (Figure 1) [34]. Arkansas schools suspended in-person learning on March 15, 2020 and many activities where children congregate during the summer were closed. We found that the seroprevalence rate in children increased modestly between spring and summer, suggesting that these protective measures effectively limited SARS-CoV-2 spread among children in Arkansas. The larger increase in seroprevalence for September/October (Wave 3), corresponded with the start of the 2020 – 2021 school year. However, masks and social distancing measures were in place, and many Arkansas students began the school year as remote learners. These actions likely reduced SARS-CoV-2 spread in schools, as reported outbreaks were relatively rare. Despite a dramatic increase in SARS-CoV-2 cases in Arkansas during November and December, the seroprevalence rate in children dropped during Wave 4 [35]. The decrease suggests that preventative measures in schools were effective at limiting SARS-CoV-2 spread among children during this time. However, seroprevalence increased sharply in Wave 5 to approximately 25%. Waves 4 and 5 were approximately three months apart and included the winter holidays, when children were outside the controlled school setting, and the corresponding nationwide surge in COVID-19 cases. Our data confirm that the surge also impacted the pediatric population, correlating with a large increase in the number of SARS-CoV-2-exposed children.

It is well appreciated that underrepresented racial and ethnic groups are disproportionately affected by the COVID-19 pandemic [36]. We found that Hispanic children were more likely to have SARS-CoV-2 antibodies compared to white children in all waves. Similarly, black children were more likely to have antibodies than white children in Wave 3 and Wave 5. The increase in SARS-CoV-2 antibodies in Hispanic and black children corresponded to an increase in antibody level in Hispanic and black adults during the same timeframe.(REF-Kennedy preprint) Higher antibody prevalence in Hispanic and black children may reflect multiple socioeconomic factors, including income inequality, economic stability, work circumstances, and housing [37]. Hispanic and black parents may be less likely to hold jobs that allow them to work remotely, which increases their potential exposure to SARS-CoV-2 in the workplace and limits their ability to utilize remote learning or in home childcare to decrease their children’s SARS-CoV-2 exposure [38, 39]. Residing with parents with a higher risk of workplace SARS-CoV-2 exposure combined with increased school/daycare attendance could drive infection rates in minority children. Our results underscore that the pandemic exacerbated existing racial and ethnic disparities.

Our data also highlight the importance of vaccinating children against SARS-CoV-2. General vaccinations in Arkansas began in January 2021 for those over 18 years and, and were expanded to 16 years and older on March 30, 2021 [40]. The CDC’s Advisory Committee on Immunization Practices recommended the Pfizer-BioNTech vaccine for 12-15 year-olds on May 10, 2021 [40]. Wave 5 samples were collected in April 2021, after the expansion of vaccinations to those 16 years and up but prior to inclusion of 12 – 15 year-olds. Only 8 of 145 total 16 – 18 year-olds in reported being vaccinated during Wave 5 and no subjects reported full vaccination at least two weeks prior to sample collection. Thus, our data reflect the pediatric seroprevalence rate in Arkansas prior to widespread vaccination efforts in children and adolescents. As authorization to vaccinate children younger than 12 years will not occur until late 2021, our findings indicate that most children in Arkansas remain susceptible to SARS-CoV-2 infection entering the 2021 – 2022 school year. Importantly, we found that the seroprevalence rate in younger children was lower than for older children and adolescents, which strongly emphasizes the ongoing risk of infection for a vulnerable part of the population that cannot yet be vaccinated. This is a critical consideration for policy makers as more infectious variants emerge that exhibit increased infection and likelihood of causing sever disease in younger portions of the population.

### Limitations

Remnant serum samples may not provide an accurate representation of the Arkansas population The strengths and limitations of convenience samples are described elsewhere [41]. As our study samples were collected from health clinics, our sampling method may favor subjects who were more ill or more willing to seek health care It is possible that our seroprevalence rates are overestimated due to potential cross reactivity between SARS-CoV-2 antigens and seasonal coronaviruses [42]. Some uninfected individuals, particularly children and adolescents, have antibodies that react with the SARS-CoV-2 Spike protein [42]. Further, we measured a combination of IgM and IgG antibody isotypes, as opposed to testing solely for IgG [43]. Although studies indicate that IgG and IgM responses to SARS-CoV-2 can develop simultaneously, some individuals develop detectable IgM and IgG antibodies at different times [44, 45]. Consequently, we may identify individuals with IgM and/or IgG, whereas other assays may only identify specimens with IgG. IgM also can be more non-specific than IgG [46]. Together, these limitations may cause an overestimation of antibody reactivity. However, inclusion of multiple SARS-CoV-2 antigens and the BSA control limit the non-specific reactions detected in our assay.

## CONCLUSIONS

Analysis of remnant samples collected from children and adolescents in Arkansas demonstrate a steady increase in SARS-CoV-2 infection during the first 8 months of the pandemic, followed by a more rapid increase to 25% by the end of April 2021. This finding is notable, as it is more than twice the number of confirmed SARS-CoV-2 diagnoses in the state on the final collection date. No obvious comorbidities were identified for seropositivity in children. Racial and ethnic disparities exist, with Hispanic and black children being at increased risk for SARS-CoV-2 infection compared to white children. We conclude that SARS-CoV-2 infections in children are more common than previously recognized. With the emergence of SARS-CoV-2 variants, recognition of long-term effects of SARS-CoV-2 even after mild or asymptomatic infections, and the lack of an authorized pediatric SARS-CoV-2 vaccine, these results highlight the importance of including children in SARS-CoV-2 public health, clinical care, and research strategies. Until vaccines are approved for those under 12 years, it is important that protective measures are enacted, particularly in settings where children are grouped together, to limit the risk of infection, super-spreader events, severe disease, long-term sequelae, and death. This is especially true in states like Arkansas with low vaccination rates, which is currently experiencing a new surge in infections and hospitalizations in younger age-groups caused by more infectious SARS-CoV-2 variants.

## Data Availability

The authors confirm that the data supporting the findings of this study are available within the article.

## ACKNOWLEDGMENTS

This research was supported by funds from the state of Arkansas through the Coronavirus Aid, Relief, and Economic Security (CARES) Act, the UAMS Time-Sensitive COVID-10 Research Award Program, the UAMS Translational Research Institute (UL1TR000039, TL1TR003109, and UL1TR003107), and the Center for Microbial Pathogenesis and Host Inflammatory Response (National Institutes of Health, National Institute for General Medical Sciences P20 GM103625).

---

**Supplemental table 1.** Co-morbidities of sampled populations in Arkansas during 5 periods of SARS-CoV-2 antibody testing from April 2, 2020 to April 29, 2021.

| Number (%)<sup>a</sup> |  |  |  |  |  |  |
| --- | --- | --- | --- | --- | --- | --- |
| Characteristic | Wave 1<br>(n = 316) | Wave 2<br>(n = 300) | Wave 3<br>(n = 594) | Wave 4<br>(n = 600) | Wave 5<br>(n = 590) | Overall<br>(N = 2400) |
| Collection dates | April 2 –<br>May 6, 2020 | June 6 –<br>August 10, 2020 | September 8 –<br>October 17, 2020 | November 7 –<br>December 17, 2020 | April 5 –<br>April 28, 2021 | April 2, 2020 –<br>April 28, 2021 |
| Respiratory viral panel within 6 mo. | NR | 34 (11.3) | 35 (5.9) | 25 (4.2) | NR | 94 (6.3) |
| Asthma | 12 (3.8) | 31 (10.3) | 68 (11.5) | 62 (10.4) | 66 (11.2) | 239 (10.0) |
| Hypertension | 16 (5.1) | 16 (5.3) | 25 (4.2) | 34 (5.7) | 20 (3.4) | 111 (4.6) |
| Obesity | 14 (4.4) | 46 (15.3) | 83 (14.0) | 109 (18.3) | 85 (14.4) | 337 (14.1) |
| Diabetes | 10 (3.2) | 15 (5.0) | 30 (5.1) | 27 (4.5) | 31 (5.3) | 113 (4.7) |
| Autoimmune disease | NR | 7 (2.3) | 14 (2.4) | 19 (3.2) | 23 (3.9) | 63 (3.0) |

<sup>a</sup> Percentages calculated from non-missing values for all periods. Missing values in Wave 1 included respiratory virus panel performed within 6 months (n = 316) and autoimmune disease (n = 316), Missing values in Wave 4 included respiratory virus panel performed within 6 months (n = 5), asthma (n = 5), hypertension (n = 5), obesity (n = 5), diabetes (n = 5), and autoimmune disease (n = 5). Missing values in Wave 5 included Respiratory viral panel within 6 months (n = 590).
NR = not reported

**Supplemental table 2.** Marginal association between SARS-CoV-2 antibody positivity and demographic characteristics by time period.

|  |  | Age category |  |  |  | Sex |  | Race/ethnicity |  |  |  | Metropolitan status |  |
| --- | --- | --- | --- | --- | --- | --- | --- | --- | --- | --- | --- | --- | --- |
| Wave number | Characteristic<sup>a</sup> | 1-4 years | 5-9 years | 10-14 years | 15-18 years | Female | Male | Black | Hispanic | Others | White | Urban | Rural |
| Wave 1<br>(n = 316) | No., (%) | 6 (10.7) | 3 (4.9) | 8 (8.9) | 8 (7.3) | 14 (8.7) | 11 (7.1) | 5 (5.5) | 3 (10.7) | 1 (5.6) | 16 (9.0) | NR | NR |
|  | RR<br>(95% CI) | 1.46<br>(0.53-4.00) | 0.67<br>(0.18-2.43) | 1.21<br>(0.47-3.10) | 1.0<br>(Ref) | 1.23<br>(0.57, 2.62) | NC | 0.61<br>(0.23-1.62) | 1.19<br>(0.37-3.83) | 0.62<br>(0.09-4.39) | 1.0<br>(Ref) | NC | NC |
|  | P-value<sup>b</sup> | 0.690 |  |  |  | 0.600 |  | 0.706 |  |  |  | NC |  |
| Wave 2<br>(n = 300) | No., (%) | 5 (15.2) | 6 (8.0) | 11 (11.2) | 7 (7.5) | 14 (9.0) | 15 (10.3) | 2 (3.8) | 9 (14.1) | 3 (12.5) | 15 (9.7) | 28 (11.2) | 1 (2.0) |
|  | RR<br>(95% CI) | 2.03<br>(0.69-5.97) | 1.07<br>(0.38-3.06) | 1.51<br>(0.61-3.72) | 1.0<br>(Ref) | 0.87<br>(0.44, 1.74) | 1.0<br>(Ref) | 0.39<br>(0.09-1.64) | 1.44<br>(0.67-3.13) | 1.28<br>(0.40-4.10) | 1.0<br>(Ref) | 5.47<br>(0.76-39.24) | 1.0<br>(Ref) |
|  | P-value<sup>b</sup> | 0.539 |  |  |  | 0.701 |  | 0.352 |  |  |  | 0.091 |  |
| Wave 3<br>(n = 594) | No., (%) | 16 (19.8) | 21 (18.8) | 29 (14.2) | 30 (15.3) | 56 (17.3) | 40 (14.8) | 20 (17.1) | 28 (29.5) | 9 (27.3) | 35 (10.7) | 76 (16.6) | 20 (14.7) |
|  | RR<br>(95% CI) | 1.29<br>(0.75-2.23) | 1.23<br>(0.74-2.03) | 0.92<br>(0.58-1.48) | 1.0<br>(Ref) | 1.17<br>(0.80-1.69) | 1.0<br>(Ref) | 1.60<br>(0.96-2.65) | 2.75<br>(1.77-4.28) | 2.55<br>(1.34-4.83) | 1.0<br>(Ref) | 1.13<br>(0.72-1.78) | 1.0<br>(Ref) |
|  | P-value<sup>b</sup> | 0.560 |  |  |  | 0.417 |  | **<0.0001** |  |  |  | 0.602 |  |
| Wave 4<br>(n = 600) | No., (%) | 6 (7.2) | 15 (12.9) | 30 (14.0) | 27 (14.8) | 46 (13.6) | 32 (12.5) | 23 (15.3) | 16 (20.8) | 1 (2.9) | 36 (11.5) | 55 (13.0) | 23 (13.5) |
|  | RR<br>(95% CI) | 0.49<br>(0.21-1.14) | 0.87<br>(0.48-1.57) | 0.95<br>(0.58-1.53) | 1.0<br>(Ref) | 1.09<br>(0.72-1.67) | 1.0<br>(Ref) | 1.33<br>(0.82-2.16) | 1.80<br>(1.06-3.07) | 0.25<br>(0.04-1.80) | 1.0<br>(Ref) | 0.96<br>(0.61-1.52) | 1.0<br>(Ref) |
|  | P-value<sup>b</sup> | 0.410 |  |  |  | 0.679 |  | 0.060 |  |  |  | 0.876 |  |
| Wave 5<br>(n = 590) | No., (%) | 12 (16.0) | 27 (19.0) | 58 (30.9) | 54 (29.5) | 90 (27.0) | 61 (23.9) | 31 (28.4) | 38 (43.7) | 15 (35.7) | 57 (17.9) | 101 (23.4) | 50 (31.9) |
|  | RR<br>(95% CI) | 0.54<br>(0.31-0.95) | 0.64<br>(0.43-0.97) | 1.05<br>(0.77-1.43) | 1.0<br>(Ref) | 1.13<br>(0.85-1.50) | 1.0<br>(Ref) | 1.59<br>(1.09-2.32) | 2.44<br>(1.74-3.41) | 1.99<br>(1.25-3.18) | 1.0<br>(Ref) | 0.74<br>(0.55-0.98) | 1.0<br>(Ref) |
|  | P-value<sup>b</sup> | 0.018 |  |  |  | 0.395 |  | **<0.0001** |  |  |  | 0.035 |  |

<sup>a</sup> Missing values from Supplemental table 1 were not included in the analysis.
<sup>b</sup> P-value indicates whether the positivity differs between the category of a respective variable.
Statistically significant values are shown in bold text.

**Supplemental table 3.** Marginal association between SARS-CoV-2 antibody positivity and comorbidities by time period.

|  |  | Respiratory virus panel within 6 months |  | Asthma |  | Hypertension |  | Obesity |  | Diabetes |  | Autoimmune disease |  |
| --- | --- | --- | --- | --- | --- | --- | --- | --- | --- | --- | --- | --- | --- |
| Wave number | Characteristic<sup>a</sup> | Yes | No | Yes | No | Yes | No | Yes | No | Yes | No | Yes | No |
| Wave 1<br>(n = 316) | No., (%) | NR | NR | 4 (33.3) | 21 (6.9) | 0 | 25 (8.3) | 1 (7.1) | 24 (8.0) | 3 (30.0) | 22 (7.2) | NR | NR |
|  | RR<br>(95% CI) | ND | ND | 4.83<br>(1.96-11.87) | 1.0<br>(Ref) | ND | ND | 0.90<br>(0.13-6.18) | 1.0<br>(Ref) | 4.17<br>(1.49-11.67) | 1.0<br>(Ref) | ND | ND |
|  | P-value<sup>b</sup> | ND |  | **0.003** |  | ND |  | 0.914 |  | **0.007** |  | ND |  |
| Wave 2<br>(n = 300) | No., (%) | 6 (17.7) | 23 (8.7) | 5 (16.1) | 24 (8.9) | 1 (6.3) | 28 (9.9) | 5 (10.9) | 24 (9.5) | 0 (0.0) | 29 (10.2) | 1 (14.3) | 28 (9.6) |
|  | RR<br>(95% CI) | 2.04<br>(0.89-4.65) | 1.0<br>(Ref) | 1.81<br>(0.74-4.40) | 1.0<br>(Ref) | 0.63<br>(0.09-4.37) | 1.0<br>(Ref) | 1.15<br>(0.46-2.86) | 1.0<br>(Ref) | NC | 1.0<br>(Ref) | 1.49<br>(0.24-9.49) | 1.0<br>(Ref) |
|  | P-value<sup>b</sup> | 0.090 |  | 0.192 |  | 0.643 |  | 0.763 |  | NC |  | 0.670 |  |
| Wave 3<br>(n = 594) | No., (%) | 7 (20.0) | 89 (15.9) | 6 (8.8) | 90 (17.1) | 6 (24.0) | 90 (15.8) | 15 (18.1) | 81 (15.9) | 5 (16.7) | 91 (16.1) | 2 (14.3) | 94 (16.2) |
|  | RR<br>(95% CI) | 1.26<br>(0.63-2.50) | 1.0<br>(Ref) | 0.52<br>(0.23-1.13) | 1.0<br>(Ref) | 1.52<br>(0.74-3.13) | 1.0<br>(Ref) | 1.14<br>(0.69-1.88) | 1.0<br>(Ref) | 1.03<br>(0.45-2.35) | 1.0<br>(Ref) | 0.88<br>(0.24-3.22) | 1.0<br>(Ref) |
|  | P-value<sup>b</sup> | 0.517 |  | 0.099 |  | 0.258 |  | 0.607 |  | 0.938 |  | 0.849 |  |
| Wave 4<br>(n = 600) | No., (%) | 0 (0.0) | 78 (13.7) | 5 (8.1) | 73 (13.7) | 5 (14.7) | 73 (13.0) | 16 (14.7) | 62 (12.8) | 2 (7.4) | 76 (13.4) | 2 (10.5) | 76 (13.2) |
|  | RR<br>(95% CI) | ND | ND | 0.59<br>(0.25-1.40) | 1.0<br>(Ref) | 1.13<br>(0.49-2.61) | 1.0<br>(Ref) | 1.15<br>(0.69-1.91) | 1.0<br>(Ref) | 0.55<br>(0.14-2.14) | 1.0<br>(Ref) | 0.80<br>(0.21-3.01) | 1.0<br>(Ref) |
|  | P-value<sup>b</sup> | ND |  | 0.231 |  | 0.775 |  | 0.589 |  | 0.391 |  | 0.739 |  |
| Wave 5<br>(n = 590) | No., (%) | NR | NR | 16 (24.2) | 136 (26.0) | 7 (35.0) | 145 (25.4) | 20 (23.5) | 132 (26.1) | 12 (38.7) | 140 (25.0) | 7 (30.4) | 145 (25.6) |
|  | RR<br>(95% CI) | ND | ND | 0.93<br>(0.60-1.47) | 1.0<br>(Ref) | 1.38<br>(0.74-2.54) | 1.0<br>(Ref) | 0.90<br>(0.60-1.36) | 1.0<br>(Ref) | 1.55<br>(0.97-2.46) | 1.0<br>(Ref) | 1.19<br>(0.63-2.24) | 1.0<br>(Ref) |
|  | P-value<sup>b</sup> | ND |  | 0.767 |  | 0.308 |  | 0.616 |  | 0.067 |  | 0.590 |  |

<sup>a</sup>Missing values from Supplemental table 1 were not included in the analysis.470 <sup>b</sup>P-value indicates whether the positivity differs between the category of a respective variable.
Statistically significant values are shown in bold text.
NR = not reported; ND = not determined

**Supplemental table 4.** PCR test history and results from sampled populations in Arkansas during 5 periods of SARS-CoV-2 antibody testing from April 2, 2020 to April 28, 2021.

| Number (%)<sup>a</sup> |  |  |  |  |  |  |
| --- | --- | --- | --- | --- | --- | --- |
| Characteristic | Wave 1<br>(n = 316) | Wave 2<br>(n = 300) | Wave 3<br>(n = 594) | Wave 4<br>(n = 600) | Wave 5<br>(n = 590) | Overall<br>(N = 2400) |
| Collection dates | April 2 –<br>May 6, 2020 | June 6 –<br>August 10, 2020 | September 8 –<br>October 17, 2020 | November 7 –<br>December 17, 2020 | April 5 –<br>April 28, 2021 | April 2, 2020 –<br>April 28, 2021 |
| SARS-CoV-2 RT-PCR test administered |  |  |  |  |  |  |
| Yes | 64 (20.3) | 82 (27.3) | 160 (26.9) | 173 (29.1) | 235 (39.8) | 714 (29.8) |
| No | 252 (79.7) | 218 (72.7) | 434 (73.1) | 422 (70.9) | 355 (60.2) | 1681 (70.2) |
| SARS-CoV-2 RT-PCR test result |  |  |  |  |  |  |
| Positive | 0 (0.0) | 1 (1.2) | 6 (3.8) | 11 (6.4) | 20 (8.6) | 38 (5.3) |
| Negative | 64 (100) | 81 (98.8) | 154 (96.2) | 162 (93.6) | 214 (91.5) | 675 (94.7) |

<sup>a</sup> Percentages calculated from non-missing values for all periods. Missing values in Wave 1 included SARS-CoV-2 RT-PCR test result (n = 252). Missing values in Wave 2 included SARS-CoV-2 RT- PCR test result (n = 218). Missing values in Wave 3 included SARS-CoV-2 RT-PCR test result (n = 434). Missing values in Wave 4 included SARS-CoV-2 RT-PCR test administered (n = 5) and SARS-CoV-2 RT-PCR test result (n = 427). Missing values in Wave 5 included SARS-CoV-2 PCR test result (n = 356).

**Supplemental table 5.**
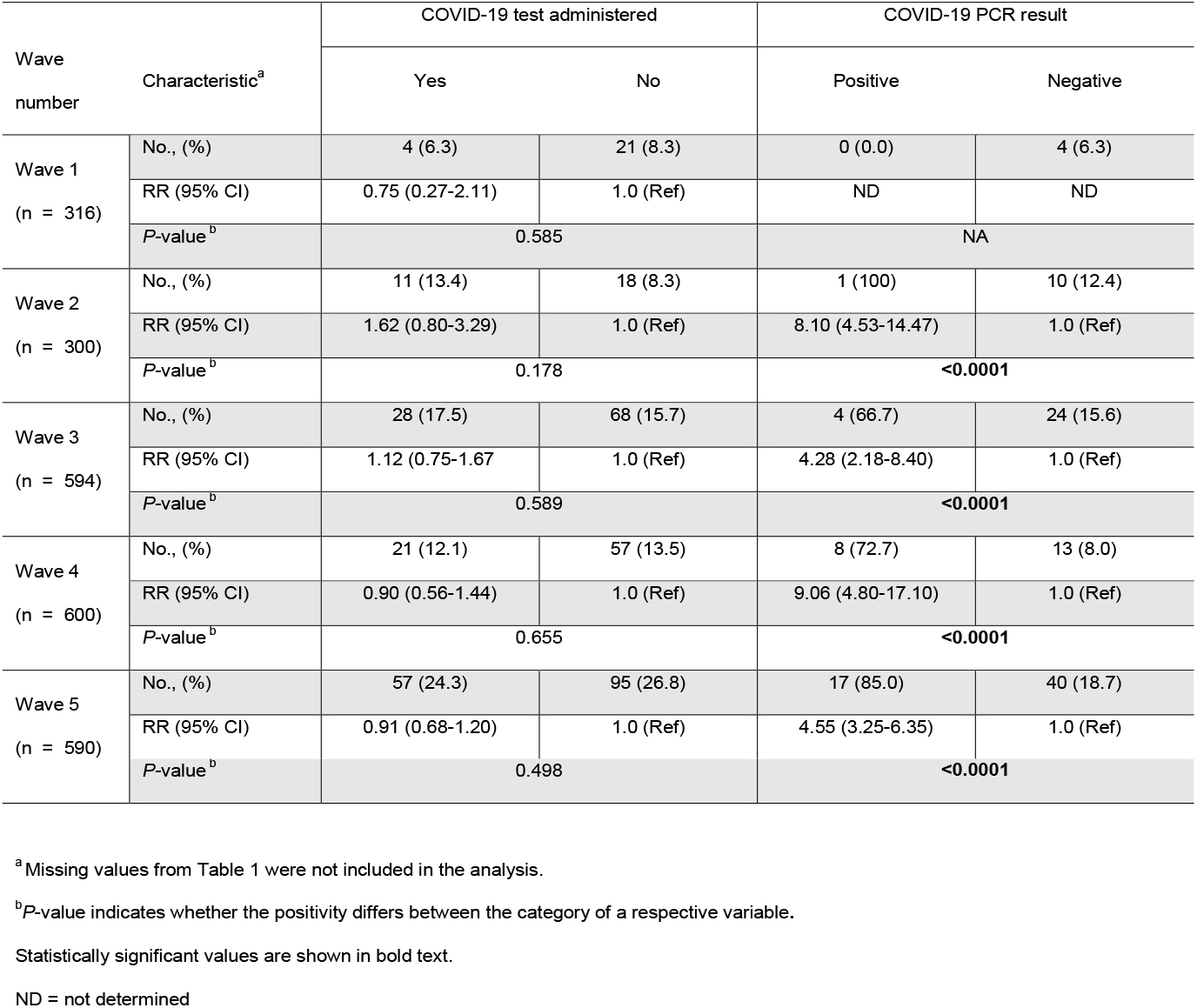
Marginal association between SARS-CoV-2 antibody positivity and SARS-CoV-2 PCR testing by time period. RT-PCR testing by time period.

| Wave number | Characteristic<sup>a</sup> | COVID-19 test administered |  | COVID-19 PCR result |  |
| --- | --- | --- | --- | --- | --- |
|  |  | Yes | No | Positive | Negative |
| Wave 1<br>(n = 316) | No., (%) | 4 (6.3) | 21 (8.3) | 0 (0.0) | 4 (6.3) |
|  | RR (95% CI) | 0.75 (0.27-2.11) | 1.0 (Ref) | ND | ND |
|  | P-value<sup>b</sup> | 0.585 |  | NA |  |
| Wave 2<br>(n = 300) | No., (%) | 11 (13.4) | 18 (8.3) | 1 (100) | 10 (12.4) |
|  | RR (95% CI) | 1.62 (0.80-3.29) | 1.0 (Ref) | 8.10 (4.53-14.47) | 1.0 (Ref) |
|  | P-value<sup>b</sup> | 0.178 |  | **<0.0001** |  |
| Wave 3<br>(n = 594) | No., (%) | 28 (17.5) | 68 (15.7) | 4 (66.7) | 24 (15.6) |
|  | RR (95% CI) | 1.12 (0.75-1.67) | 1.0 (Ref) | 4.28 (2.18-8.40) | 1.0 (Ref) |
|  | P-value<sup>b</sup> | 0.589 |  | **<0.0001** |  |
| Wave 4<br>(n = 600) | No., (%) | 21 (12.1) | 57 (13.5) | 8 (72.7) | 13 (8.0) |
|  | RR (95% CI) | 0.90 (0.56-1.44) | 1.0 (Ref) | 9.06 (4.80-17.10) | 1.0 (Ref) |
|  | P-value<sup>b</sup> | 0.655 |  | **<0.0001** |  |
| Wave 5<br>(n = 590) | No., (%) | 57 (24.3) | 95 (26.8) | 17 (85.0) | 40 (18.7) |
|  | RR (95% CI) | 0.91 (0.68-1.20) | 1.0 (Ref) | 4.55 (3.25-6.35) | 1.0 (Ref) |
|  | P-value<sup>b</sup> | 0.498 |  | **<0.0001** |  |

<sup>a</sup>Missing values from Table 1 were not included in the analysis.
<sup>b</sup>P-value indicates whether the positivity differs between the category of a respective variable.
Statistically significant values are shown in bold text.
ND = not determined

